# Effect of Endovascular therapy in large anterior circulation ischemic strokes -a systematic review and meta-analysis

**DOI:** 10.1101/2023.06.02.23290913

**Authors:** Baikuntha Panigrahi, Subhangi Thakur Hameer, Rohit Bhatia, Partha Haldar, Agrata Sharma, M.V.Padma Srivastava

## Abstract

**Background and purpose:** The benefit of endovascular treatment in large anterior circulation ischemic strokes with low ASPECTS score(<6) is uncertain. Recent studies have demonstrated the benefit of endovascular treatment (EVT) in large ischemic strokes. The present meta-analysis aims to assess the combined effect of these studies on efficacy and safety of endovascular treatment in this group of patients.

**Materials and Methods:** We conducted a systematic review and meta-analysis according to the Preferred Reporting Items for Systematic Reviews and Meta-Analyses statement. Databases MEDLINE, PubMed, EMBASE, SCOPUS, Google Scholar, Tripdatabase were searched for randomised controlled trials with at least 50 participants. The primary efficacy outcome analysed was the relative risk of functional independence defined as mRS-0-2 at 90 days. Secondary efficacy outcomes included early neurological improvement, death due to any cause at 90 days and proportion of patients requiring decompressive hemicraniectomy. The primary safety outcome was the risk of developing symptomatic intracerebral haemorrhage.

**Results:** A total of 3 studies(RESCUE Japan-LIMIT,SELECT 2 and ANGEL ASPECTS) involving 1011 patients ; 510 in the EVT arm and 501 in the medical management (MM) arm met the defined criteria (ASPECTS-3-5). The combined RR for the primary outcome of mRS 0-2 was 2.53 [1.84-3.47] (p=<0.0001) favouring EVT over MM. The primary safety outcome of symptomatic intracerebral haemorrhage was not significant in the EVT arm with a combined RR of 1.84 [0.94-3.60] (p=0.5157). Mortality rates were similar in both arms (26.67% in EVT arm vs 27.94% in MM arm) with a combined RR of 0.95 [0.78; 1.16] (p=1.000).

**Conclusion:** In patients with LVO and low ASPECTS (3-5), EVT was associated with higher likelihood of achieving functional independence and early neurologic improvement but did not provide any mortality benefit. The rates of symptomatic ICH were similar in both the groups whereas the risk of any ICH was significantly higher in the EVT arm.

**Funding-Nil:** The protocol for the review was registered with the International Prospective Register of Systematic Reviews (PROSPERO) database(CRD42023400675)

## Background and Purpose

Endovascular therapy is currently the standard of care in patients with acute ischemic stroke due to large vessel occlusion (LVO) and can be considered up to 24 hours (in selected patients) from the onset of symptoms. The ACC (American College of Cardiology) /AHA (American Heart Association) 2018 guidelines recommend mechanical thrombectomy with a stent retriever or direct aspiration in the 0-6 h window period if patients meet defined criteria including ASPECTS of ≥ 6.^1^ In the 6-24 hours window, endovascular therapy is recommended in patients meeting the DAWN ^2^ or DEFUSE3 ^3^ eligibility criteria.

The benefit and safety of endovascular therapy (EVT) in patients with acute ischemic strokes in the anterior circulation with a large infarct (ASPECTS<6) is an important knowledge gap in current stroke management guidelines. Such patients are usually managed conservatively and decompressive surgery is advised if they deteriorate clinically or radiologically.^4^ The futility of EVT in these cases is based on assumptions rather than evidence as most of these patients have traditionally been excluded from trials of EVT. There has been an interest in offering EVT in large strokes after observational studies showed benefit of EVT in the ASPECTS 3-5 group. The present systematic review assesses the current evidence from randomised controlled trials on the efficacy of EVT in patients with large anterior circulation strokes. The objective of the study in the PICO format is summarised in Table 1.

**Table 1.** The Research Question in PICO format

| PICO Statements |  |
| --- | --- |
| Patients | Patients of age $\geq 18$ years with a baseline mRS-0-1 who had an acute ischemic stroke within 24hrs of last known well with a NIHSS $\geq 6$, strokes involving the anterior circulation, large infarct core defined as an Alberta Stroke Program Early Computed Tomography (ASPECT) Score of 5 or below or a core volume of $\geq 70$ ml on computed tomography perfusion or diffusion-weighted magnetic resonance imaging. |
| Intervention | Endovascular Therapy with any device as per the discretion of the treating neuro-interventionist. |
| Control | Medical Management. |
| Outcomes | The primary efficacy outcome was the relative risk of functional independence defined as mRS- 0-2 at 90 days.<br>Secondary efficacy outcomes included Early neurological improvement (ENI) defined as a decrease in NIHSS by 8 points or more or 0-1 if NIHSS $\leq 8$ at 24-36 hrs, death due to any cause at 90 days and proportion of patients requiring decompressive hemicraniectomy at any time post the intervention or on standard medical care.<br>The primary safety outcome studied was the risk of developing symptomatic ICH defined as per the respective study definition. Other explanatory variables studied were the rates of systemic thrombolysis used and effect on outcomes. |

## Materials and methods

### Overview

We used the PRISMA (Preferred Reporting Items for Systematic Reviews and Meta-Analyses) statement for reporting systematic reviews and meta-analyses as a guide for this study. The protocol for the review was registered with the International Prospective Register of Systematic Reviews (PROSPERO) database. (CRD42023400675)

### Selection Criteria

Inclusion criteria of the studies included randomised controlled trials with a minimum sample size of 50, age >18 years, baseline mRS-0-1, acute ischemic stroke within 24hrs of last known well, NIHSS > 6, strokes involving the anterior circulation, large infarct core defined as an Alberta Stroke Program Early Computed Tomography (ASPECT) Score of 5 or below or a core volume of >70 ml on computed tomography perfusion or diffusion-weighted magnetic resonance imaging, use of endovascular therapy with any device with or without bridging IV Thrombolytic agent, articles written in English Exclusion criteria for the studies included observational cohort studies, case series, unavailable primary efficacy and safety outcome data [modified Rankin scale at 90 days and rates of symptomatic Intracranial haemorrhage(sICH)]

### Data Extraction

A comprehensive preliminary search was performed by 2 authors (B.P and S.TH) independently in the MEDLINE, PubMed, EMBASE, SCOPUS, Google Scholar, Tripdatabase and the Cochrane databases for the identification of published literature on the topic and also to confirm the absence of any published previous systematic reviews and meta-analyses, and any ongoing systematic review and meta-analysis on the same topic in PROSPERO using the following MeSH terms-Endovascular treatment, Mechanical Thrombectomy, Endovascular therapy, Revascularisation, Reperfusion, Ischemic Stroke, Acute, Ischaemic Stroke, Large Vessel Occlusion, ASPECTS, Large core, modified Rankin Scale with filters to include only Randomised Controlled Trials (RCT). Duplicates were removed after identification. Titles and abstracts were reviewed independently by 2 authors (B.P. and S.TH) to evaluate for inclusion and screening of full text. Any discrepancy was resolved by the third author (R.B). Full texts were retrieved for further consideration for inclusion in the study.

### Outcomes

The primary efficacy outcome extracted was the relative risk of functional independence defined as mRS-0-2 at 90 days. Secondary efficacy outcomes extracted were Early neurological improvement (ENI) defined as a decrease in NIHSS by 8 points or more or 0-1 if NIHSS < 8 at 24-36 hrs, death due to any cause at 90 days and the proportion of patients requiring decompressive hemicraniectomy at any time post the intervention or on standard medical care. The primary safety outcome extracted was the risk of developing symptomatic ICH defined as per the respective study definition. Other explanatory variables studied were the rates of systemic thrombolysis used and effect on outcomes.

### Assessment of Risk of Bias

After the selection of the relevant studies, the data was extracted on MS Excel by 2 authors independently (B.P. and S.TH). The information which was extracted included: the first author of the study, year of publication, number of patients included in the study, age at diagnosis, baseline characteristics of the patients included, mRS outcomes at 3 months, NIHSS at 24hrs, risk of symptomatic ICH, need for decompressive surgery and mortality. Any discrepancy among the observers was resolved in consultation with the third author (R.B.). The ROB2 tool was used to assess the risk of bias of the studies included.

### Statistical Analysis

Statistical analyses were performed using R project for statistical computing ;https://www.r-project.org/. Both fixed and random-effects models were used in view of expected heterogeneity to be present due to known differences in stroke patient characteristics and symptom definitions.

The overall effect estimate and 95% CI were used to generate Forrest plots. Statistical heterogeneity was assessed by the Higgin I^2^ statistics.

We considered study-level estimates to be heterogeneous if the I^2^ value was >50% I^2^ from 50% to 75% was considered as substantial heterogeneity, and I^2^ >75% was indicated as considerable heterogeneity.

We did not explore further possible causes of heterogeneity among study results and also we did not do subgroup analysis or meta-regression due to limited number of studies and data variables.

We conducted a sensitivity analyses to assess robustness of the synthesized results by performing leave one study out analysis and presenting a consolidated result for the primary outcome.

Certainty in the body of evidence was judged using the GRADE CRITERIA (https://www.gradeworkinggroup.org/).

## Results

### Study selection

A total of 613 studies were identified and 234 duplicates were removed. 379 studies were screened for eligibility of which 341 were excluded as 14 were in languages other than English, 73 were Retrospective studies, 58 were systematic reviews and meta-analysis and 196 studies did not meet inclusion criteria. 38 studies underwent full text screening and a total of three studies involving 1011 patients ; 510 in endovascular therapy (EVT) arm and 501 in medical management (MM) arm met the defined inclusion and exclusion criteria. All the included were RCTs. ^5,6,7^ The details are outlined in the CONSORT diagram (Figure 1).

**Figure 1.**
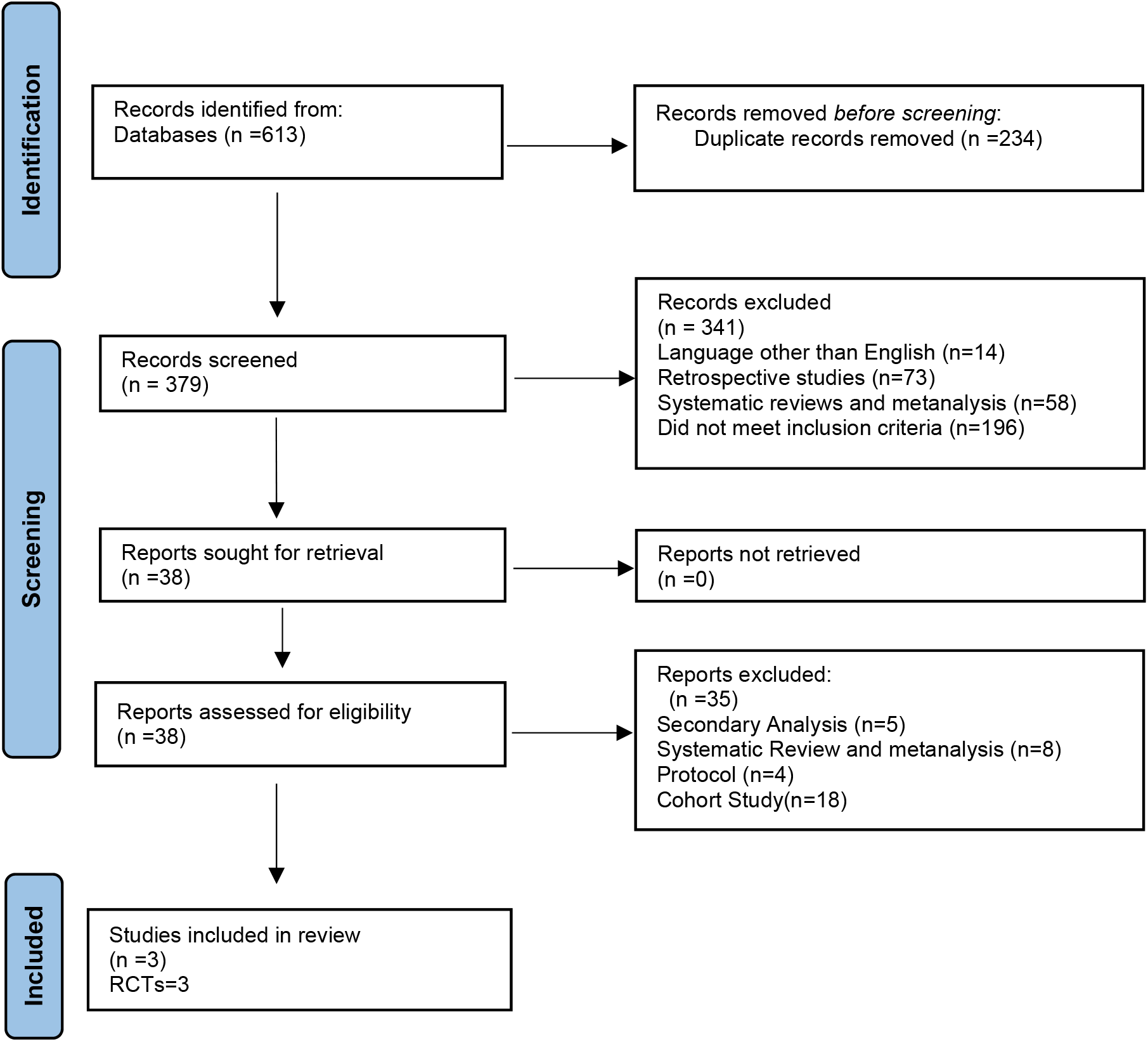
PRISMA Guidelines.

### Study characteristics

The baseline characteristics of the patients included in the studies are summarised in Table 2. No significant differences in the baseline characteristics were seen among the EVT and MM arms in the studies included. The rates of IV thrombolysis ranged from 17.2-28.6% among patients included across groups.

**Table 2.** Baseline characteristics of the patients included

| Baseline characteristics | RESCUE-Japan<br>LIMIT |  | SELECT 2 |  | ANGEL-ASPECTS |  |
| --- | --- | --- | --- | --- | --- | --- |
| Total Sample size | 203 |  | 352 |  | 456 |  |
| Age in years | Mean (SD)=76.6<br>(10) |  | Median(IQR)=66(58-75) |  | Median(IQR)=68(61-73) |  |
|  | EVT | MM | EVT | MM | EVT | MM |
| n | 101 | 102 | 178 | 174 | 231 | 225 |
| Males (%) | 55<br>(54.4) | 58<br>(56.9) | 107 (60.1) | 100<br>(57.5) | 135 (58.4) | 144 (64) |
| Hypertension (%) | 71<br>(70.3) | 70<br>(68.6) | 136 (76.4) | 134 (77) | 136 (58.9) | 136<br>(60.4) |
| Smokers (%) | 18<br>(17.8) | 21<br>(20.6) |  |  | 67 (29) | 77 (34.2) |
| Diabetes (%) | 23<br>(22.8) | 20<br>(19.6) | 53 (29.8) | 55 (31.6) | 43 (18.6) | 40 (17.8) |
| Atrial Fibrillation (%) | 60<br>(59.4) | 60<br>(58.8) | 47 (26.4) | 38 (21.8) | 57(24.7) | 47 (20.9) |
| Prior Strokes (%) | 25<br>(24.8) | 26<br>(25.5) | 19 (10.7) | 13 (7.5) | 37 (16) | 36 (16) |
| Median mRS prior | 0 | 0 |  |  | 0 | 0 |
| Median NIHSS | 22 | 22 | 19 | 19 | 16 | 15 |
| Site of Occlusion |  |  |  |  |  |  |
| ICA | 47 | 49 | 80 | 66 | 83 | 81 |
| M1 | 74 | 70 | 91 | 100 | 145 | 142 |
| M2 | 0 | 3 | 7 | 8 | 2 | 2 |
| Tandem | 20 | 20 | 56 | 44 | 41 | 35 |
| ASPECTS (0-2) | 5 | 3 | 12 | 8 | 32 | 30 |
| ASPECTS (3-5) | 96 | 99 | 148 | 142 | 198 | 195 |
| CTP/DWI >70mL core |  |  | 179 | 171 | 90 | 96 |
| Median Core Volume mL | 94 | 110 | 81.5 | 79 | 60.5 | 63 |
| Median Duration between LKW<br>and hospital arrival in min | 190 | 170 | 587.4 | 544.2 | 338 | 331 |
| IV Thrombolysis (%) | 27<br>(26.7) | 29<br>(28.4) | 37 (20.8) | 30 (17.2) | 66 (28.6) | 63 (28) |

EVT- endovascular Treatment,MM- Medical management , mRS – modified Rankin scale ,ICA- internal carotid Artery ,M1- middle cerebral Artery M1 segment ,M2- Middle Segment Artery M2 segment, ASPECTS - Alberta stroke program early CT score ,CTP- CT perfusion , DWI- diffusion Weighted imaging , LKW- Last Known Well

**Table 3.**
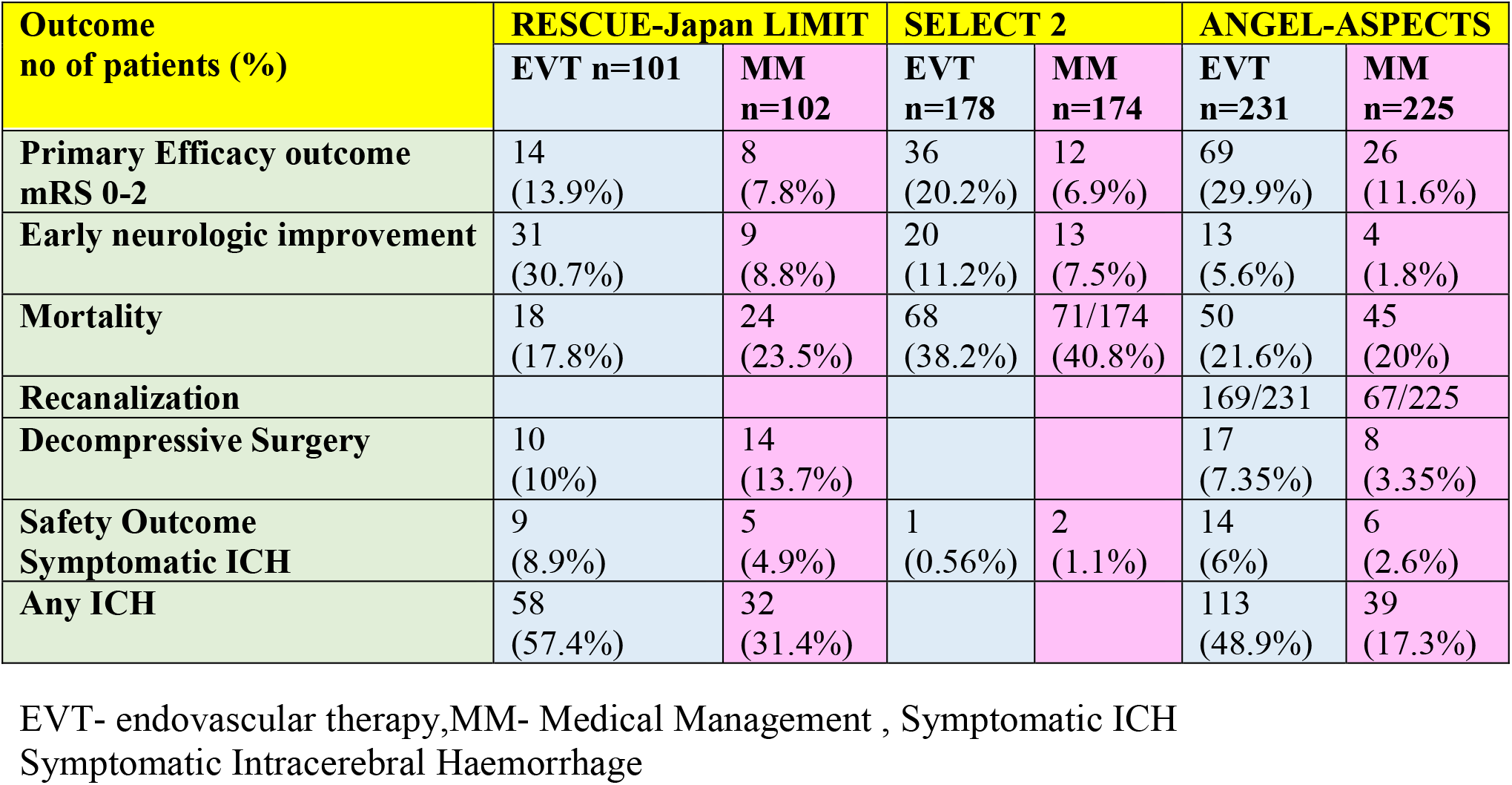
Outcomes of the three trials included

| Outcome<br>no of patients (%) | RESCUE-Japan LIMIT |  | SELECT 2 |  | ANGEL-ASPECTS |  |
| --- | --- | --- | --- | --- | --- | --- |
|  | EVT n=101 | MM n=102 | EVT n=178 | MM n=174 | EVT n=231 | MM n=225 |
| **Primary Efficacy outcome**<br>**mRS 0-2** | 14<br>(13.9%) | 8<br>(7.8%) | 36<br>(20.2%) | 12<br>(6.9%) | 69<br>(29.9%) | 26<br>(11.6%) |
| **Early neurologic improvement** | 31<br>(30.7%) | 9<br>(8.8%) | 20<br>(11.2%) | 13<br>(7.5%) | 13<br>(5.6%) | 4<br>(1.8%) |
| **Mortality** | 18<br>(17.8%) | 24<br>(23.5%) | 68<br>(38.2%) | 71/174<br>(40.8%) | 50<br>(21.6%) | 45<br>(20%) |
| **Recanalization** |  |  |  |  | 169/231 | 67/225 |
| **Decompressive Surgery** | 10<br>(10%) | 14<br>(13.7%) |  |  | 17<br>(7.35%) | 8<br>(3.35%) |
| **Safety Outcome**<br>**Symptomatic ICH** | 9<br>(8.9%) | 5<br>(4.9%) | 1<br>(0.56%) | 2<br>(1.1%) | 14<br>(6%) | 6<br>(2.6%) |
| **Any ICH** | 58<br>(57.4%) | 32<br>(31.4%) |  |  | 113<br>(48.9%) | 39<br>(17.3%) |

EVT- endovascular therapy,MM- Medical Management , Symptomatic ICH Symptomatic Intracerebral Haemorrhage

**Table 4.**
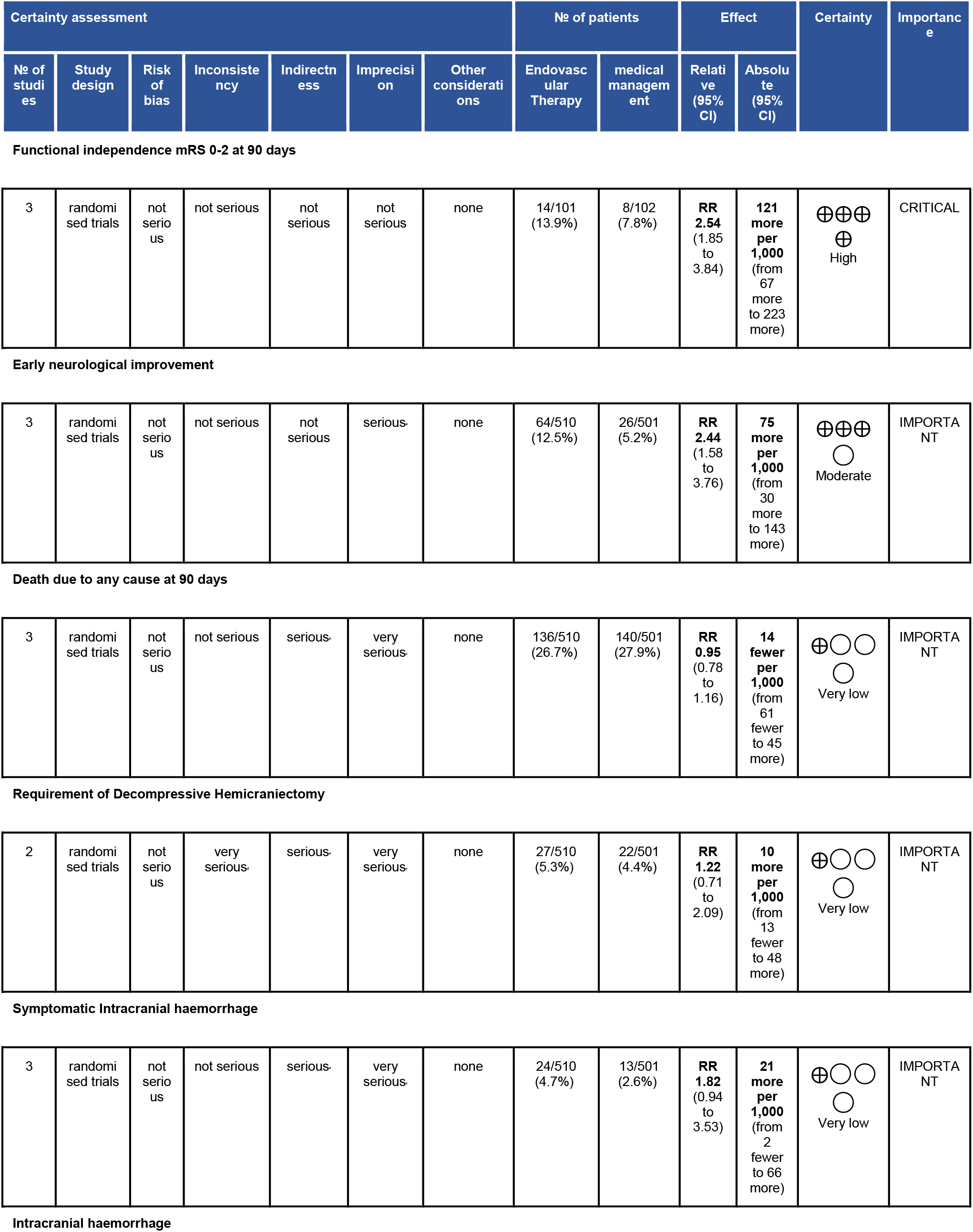
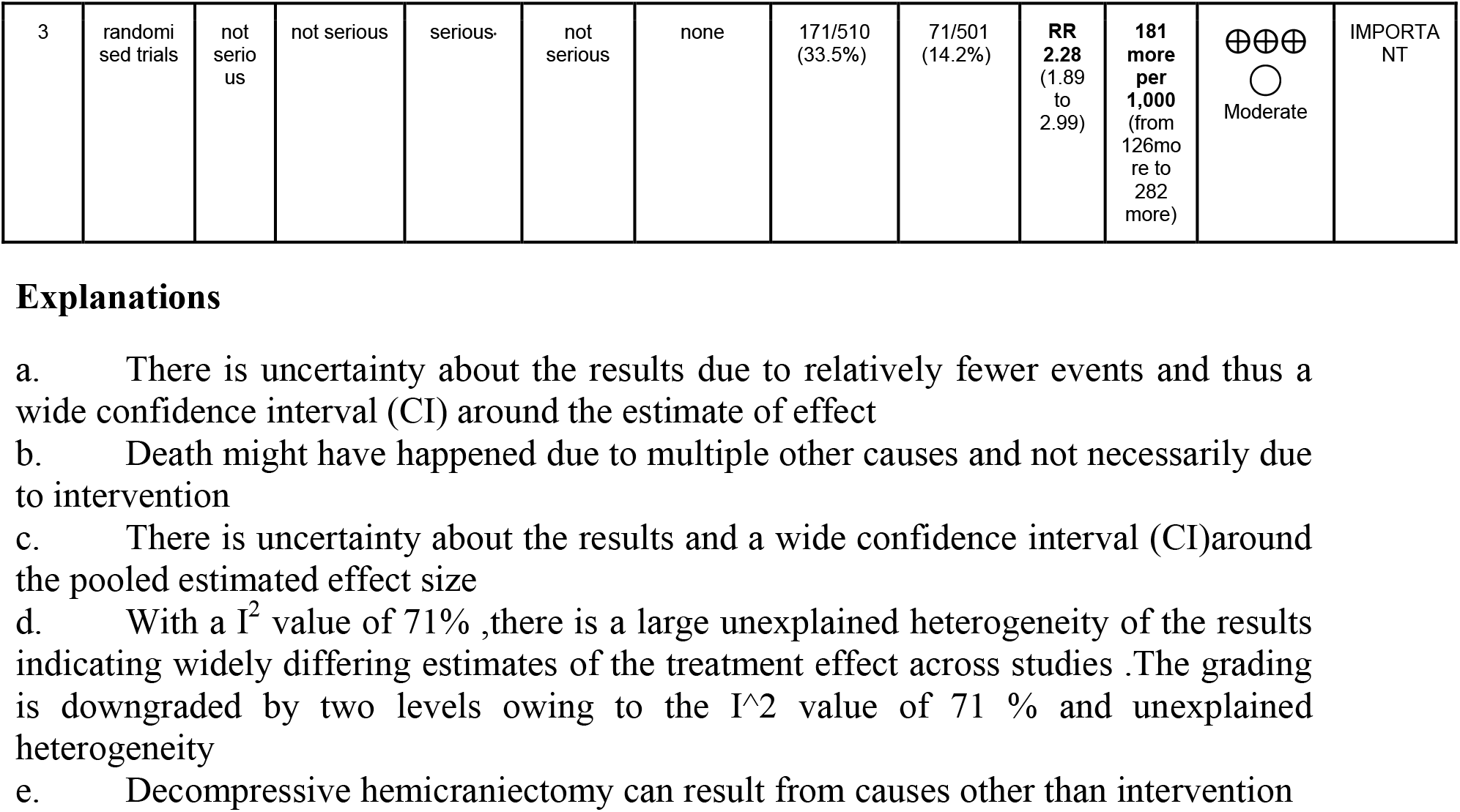
GRADE Tool for Certainty of Evidence

| Certainty assessment |  |  |  |  |  |  | Nº of patients |  | Effect |  | Certainty | Importance |
| --- | --- | --- | --- | --- | --- | --- | --- | --- | --- | --- | --- | --- |
| Nº of studies | Study design | Risk of bias | Inconsistency | Indirectness | Imprecision | Other considerations | Endovascular Therapy | medical management | Relative (95% CI) | Absolute (95% CI) |  |  |
| Functional independence mRS 0-2 at 90 days |  |  |  |  |  |  |  |  |  |  |  |  |
| 3 | randomised trials | not serious | not serious | not serious | not serious | none | 14/101 (13.9%) | 8/102 (7.8%) | RR 2.54 (1.85 to 3.84) | 121 more per 1,000 (from 67 more to 223 more) | High | CRITICAL |
| Early neurological improvement |  |  |  |  |  |  |  |  |  |  |  |  |
| 3 | randomised trials | not serious | not serious | not serious | serious | none | 64/510 (12.5%) | 26/501 (5.2%) | RR 2.44 (1.58 to 3.76) | 75 more per 1,000 (from 30 more to 143 more) | Moderate | IMPORTANT |
| Death due to any cause at 90 days |  |  |  |  |  |  |  |  |  |  |  |  |
| 3 | randomised trials | not serious | not serious | serious | very serious | none | 136/510 (26.7%) | 140/501 (27.9%) | RR 0.95 (0.78 to 1.16) | 14 fewer per 1,000 (from 61 fewer to 45 more) | Very low | IMPORTANT |
| Requirement of Decompressive Hemicraniectomy |  |  |  |  |  |  |  |  |  |  |  |  |
| 2 | randomised trials | not serious | very serious | serious | very serious | none | 27/510 (5.3%) | 22/501 (4.4%) | RR 1.22 (0.71 to 2.09) | 10 more per 1,000 (from 13 fewer to 48 more) | Very low | IMPORTANT |
| Symptomatic Intracranial haemorrhage |  |  |  |  |  |  |  |  |  |  |  |  |
| 3 | randomised trials | not serious | not serious | serious | very serious | none | 24/510 (4.7%) | 13/501 (2.6%) | RR 1.82 (0.94 to 3.53) | 21 more per 1,000 (from 2 fewer to 66 more) | Very low | IMPORTANT |
| Intracranial haemorrhage |  |  |  |  |  |  |  |  |  |  |  |  |
| 3 | randomised trials | not serious | not serious | serious | not serious | none | 171/510<br>(33.5%) | 71/501<br>(14.2%) | RR<br>2.28<br>(1.89 to 2.99) | 181 more per 1,000 (from 126 more to 282 more) | ⊕⊕⊕<br>○<br>Moderate | IMPORTANT |

### Risk of bias assessment of the included Studies

All the three trials included had a low risk of bias in the randomisation domain. There were some concerns in the risk of bias due to deviations from the intended interventions (effect adhering to intervention) as the participants were aware of the intervention and participants, carers or people delivering the interventions were aware of intervention groups.

There was low risk of bias in the outcome data as data was available for >95% of the participants in the study. The outcome assessors were blinded to the intervention in all the three trials but there is a possibility of measurement error or misclassification bias as the certification status for measurement of primary outcome was not mentioned explicitly in any of the three trials. The outcome was analysed according to a prespecified plan.

### Outcomes

### Functional independence mRS 0-2 at 90 days

The primary efficacy outcome of mRS 0-2 at 90 days was achieved in 119/510 (23.3%) patients in the EVT group and 46/501(9.18%) in the MM group (Table 2). The combined relative risk for the primary outcome utilising the fixed and random effects model were 2.54 [1.85-3.48] and 2.53 [1.84-3.47] respectively favouring EVT (p=<0.0001). (Figure-2) A mRS 0-3 at 90 days was seen in 206/510 (40.4%)patients in the EVT group and 120/501 (23.9%) patients in the MM arm (Figure 3). The combined RR for this outcome being 1.69 [1.40-2.03] using a fixed and random effects model favouring EVT (p=0.0007).

**Figure 2.**
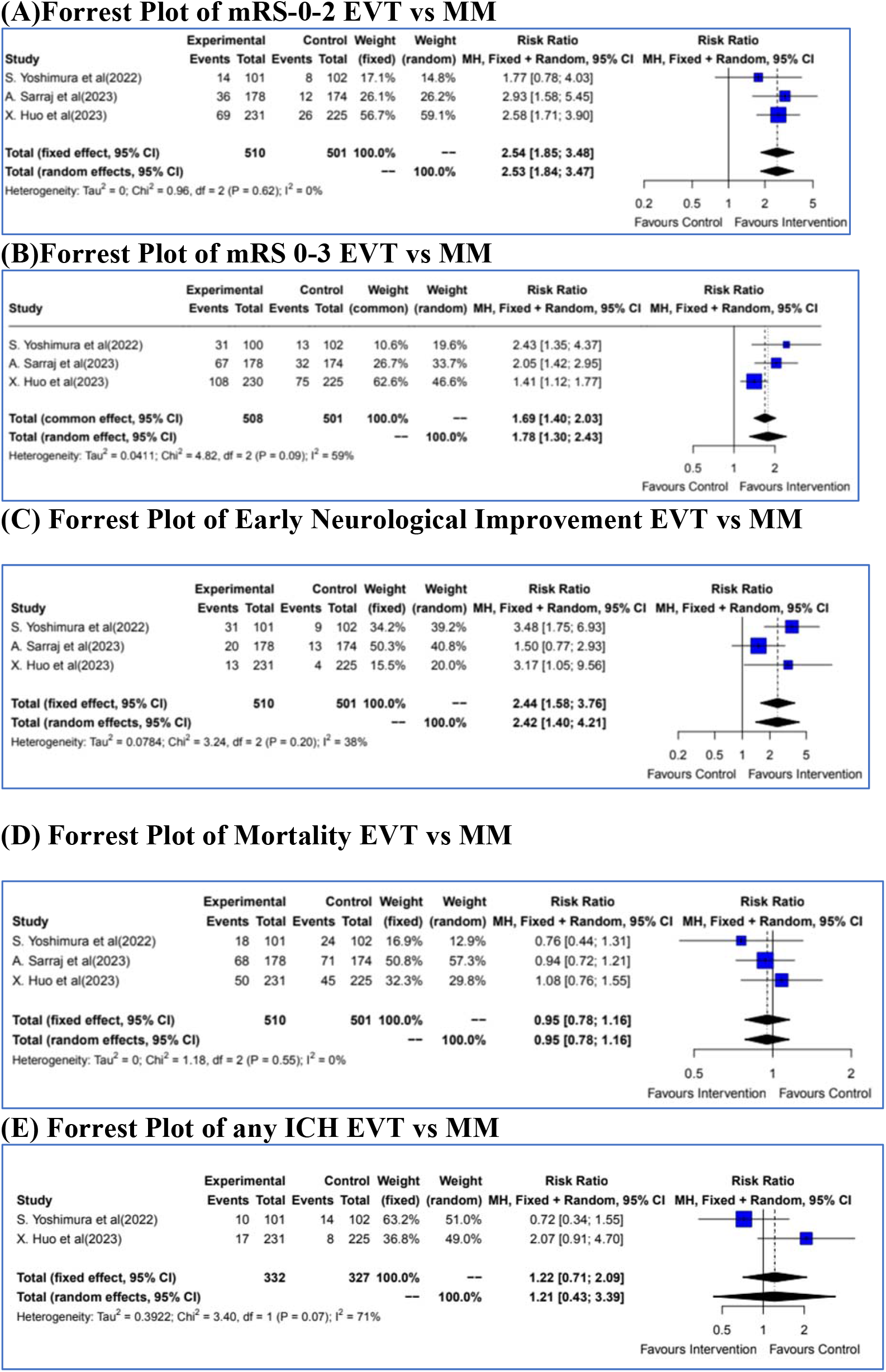
Forrest Plot of the Efficacy Outcomes.

**Figure 3.**
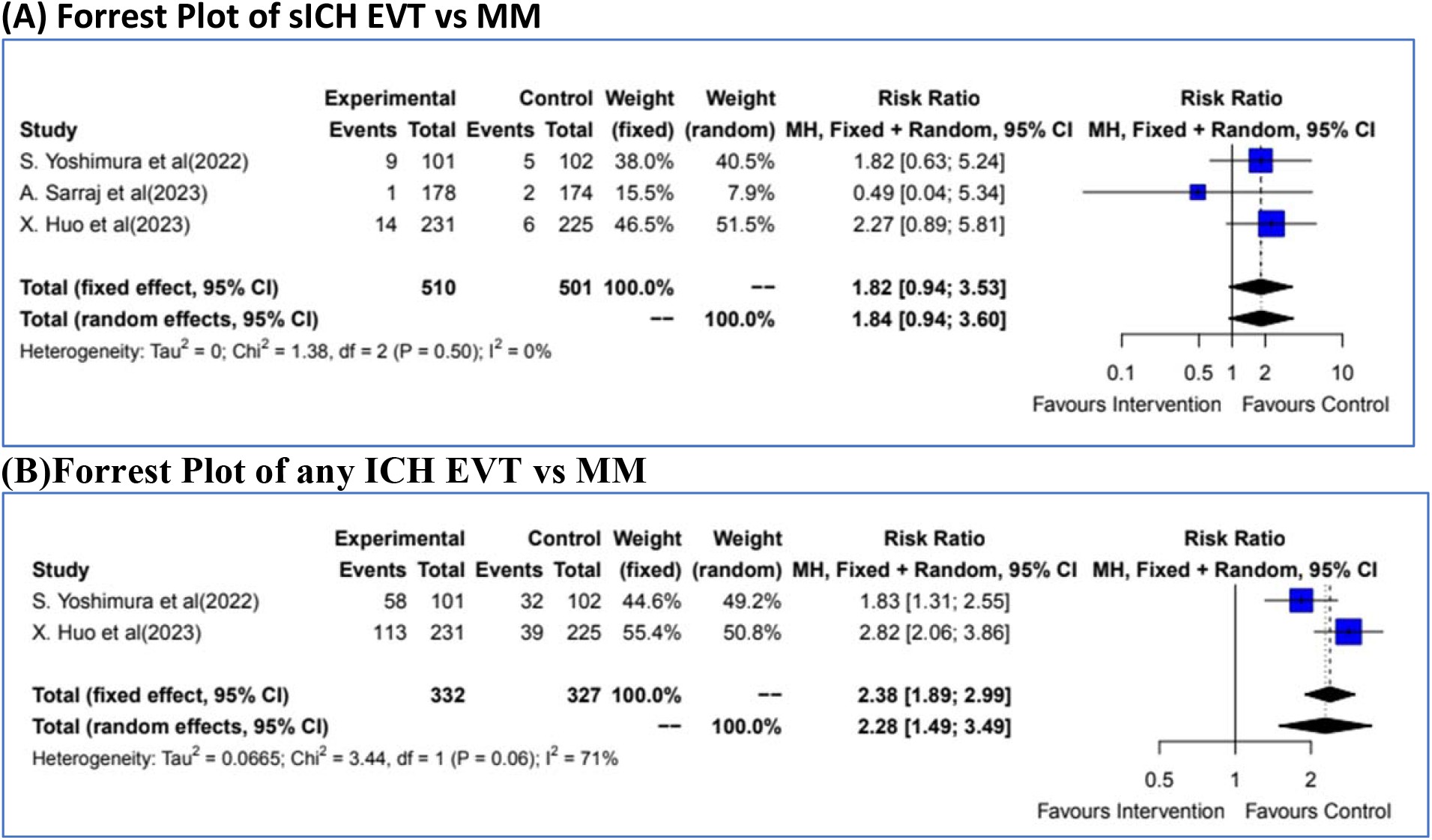
Safety outcomes.

### Early neurologic improvement (ENI)

The secondary efficacy outcome of early neurologic improvement (ENI) was observed in 64/510 (12.55%) patients in the EVT arm and in 26/501 (5.19%) patients in the MM arm (Table-2).

The combined RR for ENI utilising the fixed and random effect model was 2.44 [1.58-3.76] and 2.42 [1.40-4.21] respectively, favouring EVT. (Figure-2)

### Death due to any cause at 90 days

A total of 136/510 (26.67%) patients died in the EVT arm as compared to 140/501 (27.94%) in the MM arm with a combined RR of 0.95 [0.78; 1.16] (fixed and random effects models) (p=1.000). (Figure 2)

### Decompressive Hemicraniectomy

Decompressive surgery was required in 27/332 (8.13%) patients in the EVT arm as compared to 22/327 (6.72%) patients in the MM arm with a combined RR by fixed and random effects models of 1.22 [0.71-2.09] and 1.21 [0.43-3.39] respectively (p=1.000).

### Symptomatic ICH (Figure-3)

The primary safety outcome of symptomatic ICH was seen in 24/510 (4.7%) patients in the EVT arm as compared to 13/501 (2.6%) patients in the MM arm with a combined RR by fixed and random effects models of 1.82 [0.94-3.53] and 1.84 [0.94-3.60] respectively (p=0.5157).

Any ICH was seen in 171/332 (51.5%) patients in the EVT arm as compared to 71/327 (21.71%) patients in the MM arm with a combined RR by fixed and random effects models of 2.38 [1.89-2.99] and 2.28 [1.49-3.49] respectively (p=0.0007).

### Thrombolysis (Figure-4)

Among a subset of patients undergoing thrombolysis a combined risk ratio of benefit was 3.37 [1.54-7.41] favouring thrombolysis.

**Figure 4.**
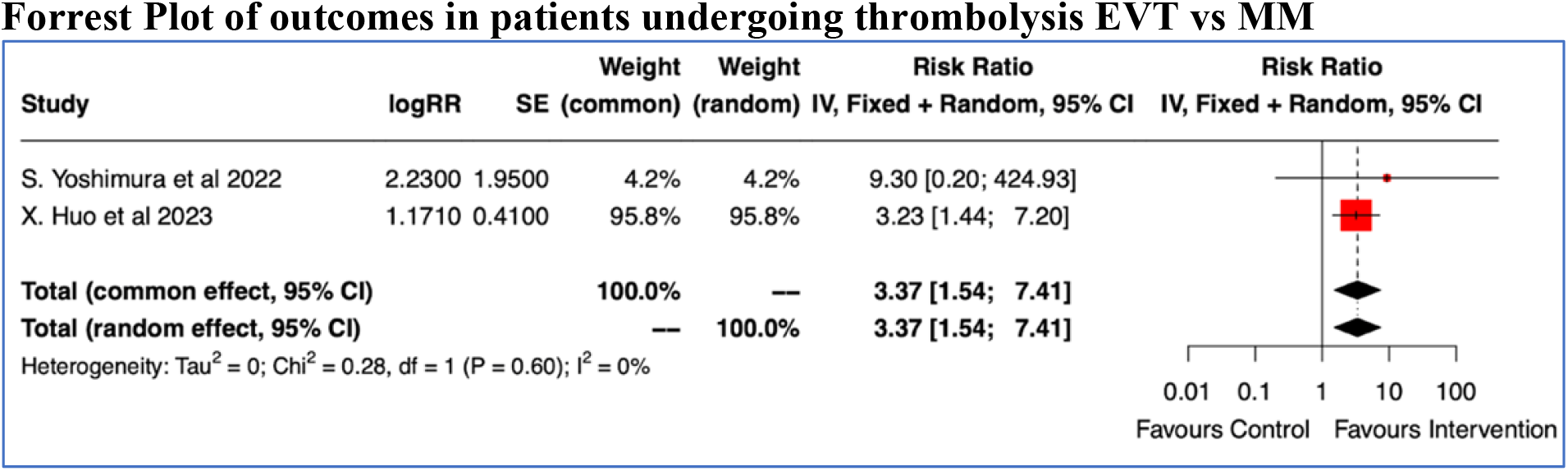
Thrombolysis and EVT outcomes.

## Discussion

This analysis provides evidence for the benefit of EVT among patients with large ischemic strokes involving the anterior circulation. Such patients undergoing EVT are 2.53 times more likely to achieve functional independence as compared to standard medical care. Considering the likelihood of relatively poor prognosis in these patients, a higher chance of achieving mRS of 0-2 is reassuring. (Figure-2). Since the pooled ARR with EVT in achieving mRS of 0-2 at three months was 14.12% yielding a NNT of 7.1 is significant considering that the HERMES collaboration demonstrated a number needed to treat (NNT) of five for achieving the same outcome. The findings are consistent with a previous meta-analysis including 17 studies and 1378 patients with an ASPECTS 0–6 and a higher odds of mRS 0–2 in the EVT arm (OR 4.76, p=0.01).^8^

The benefit of EVT even in patients with large infarcts could be due to the inherent limitations of the ASPECTS score and a strategic salvage of eloquent areas such as internal capsule and Rolandic cortex.^9^ The ASPECTS score could have inter-rater agreement concerns^11^ and may improve with lower ASPECT scores.^12,13,14^ We do not know the inter-rater agreement in the selected trials for the meta-analysis. Both anatomical and methodological factors explain why the agreement among different ASPECTS territories is so varied. Vulnerability to ischemic changes due to hypoperfusion also varies between different locations of the brain, with the insular cortex, precentral gyrus and basal ganglia being the most sensitive.

Another important predictor of the outcome is the collateral status of the patient although most patients with low ASPECTS have poor collaterals but as many as one-third may present with adequate collaterals.^15^ When collateral status was assessed in low ASPECTS patients, those with poor collaterals had a median mRS score of 5 despite successful recanalization, while patients with good collaterals showed a median mRS score of 2 after successful recanalization.^16,17^Although collateral status information was not mentioned in the trials, it is likely that patients who had a good outcome had good collaterals.

CT Perfusion also particularly in the early time window tends to overestimate the core volume. Theoretically, after sudden stoppage of blood supply to a region in the brain, the cells distal to the arterial occlusion undergo oxygen deprivation, resulting in a compensatory increase in CBF(Cerebral blood flow). Over time, these compensatory mechanisms fail, leading to a progressive decline in CBF and CBV (Cerebral blood volume), with transformation of the penumbra into irreversibly damaged tissue. Due to continued cellular oxygen deprivation, disruption of the blood brain barrier ensues, leading to the influx of ions and subsequently the net water uptake (NWU). The combined assessment of CTP parameters and NWU could reflect these stages; in patients with large CBV deficits, but low NWU, the compensatory mechanisms are largely maintained and the core lesion is likely “reversible”. On the other hand, those with large CBV lesions and high NWU have more likely crossed the threshold into irreversible tissue damage. This is known as the “ghost infarct core” phenomenon, and is particularly common in the early time window.^17^ Net water uptake (NWU) is easily and quickly assessed and when taken together, NWU and CTP-based core estimation parameters could provide a more accurate picture of both lesion size and stage of infarction at admission. In a previous study, core-overestimation was more likely to occur in patients with large perfusion cores and low net water uptake (NWU) at baseline. This could also have possibly occurred in the trials as a subgroup of patients might have a low NWU and have high cores which were overestimated and could have benefited from recanalization.^18^

Volume of the infarct in isolation may not be a good predictor of the stroke severity. The combination of volume and location may result in a significantly better correlation with clinical deficit severity (r=0.79, P=0.032).^19^ It is likely that the location of these large infarcts may have relatively spared the eloquent regions leading to a benefit of reperfusion treatment.

SELECT2 and ANGEL-ASPECTS studies were prematurely terminated and may have led to an overestimation of benefit as the target sample size was smaller than anticipated and underpowered for subgroup analyses. The benefit of EVT in patients of extremely poor ASPECTS (0-2) is still unknown as they were excluded from these studies.

Although the absolute reasons for benefit of EVT in large strokes remains elusive, further studies are needed to know which subset of these large strokes are more likely to benefit from EVT as these results could lead to a change in practice which actually renders complex imaging selection criteria difficult in acute stroke.

Changing the yardstick of outcomes changes the risk ratios as can be seen in the Forrest plots. (Figure-2) The primary outcome for the RESCUE-Japan LIMIT was mRS of 0-3 whereas the other trials used a shift analysis for the primary outcome. This is the reason why the outcome results in the mRS 0-3 are better than in mRS 0-2. An outcome of mRS 0-3 might be as bad for a large core infarct in view of the expected worse outcomes in such groups.

Safety was demonstrated in previous meta-analysis and the present study as the rates of symptomatic ICH were same in both arms although the risk of any ICH was higher in the EVT arm. In an analysis of blood pressure after EVT (BEST) multicentre prospective registry of patients treated with EVT, low ASPECTS was found to be the most important determinant of sICH. ASPECTS < 6 carried a statistically significant (p = 0.009) odds ratio of 10 for sICH post EVT. All the three present trials showed that despite the odds of sICH, there is a high likelihood of functional independence in these group of patients.^20^

Another factor with respect to the safety benefit of EVT in low ASPECTS patients could be related to the reduction of brain edema and malignant mass effect, leading to reduced rates of decompressive hemicraniectomy.^21^

Another important point to note is that even in patients with a low ASPECTS thrombolysis was associated with good outcome contrary to the assumptions. (Figure-4) Although definite conclusions cannot be drawn from the present study in view of the analysis based on the relative risks and the absolute numbers and individual patient data was not available for knowing recanalization rates the study provides some insights into safety of Intravenous Thrombolysis in this often excluded group of patients. Large scale studies are needed to conclusively prove benefit.

We conducted the analysis of the evidence for the outcomes using the GRADE methodology .It showed that the certainty of the intervention i.e EVT in low ASPECTS leading to functional independence mRS 0-2 at 90 days is high.

The Limitation of this study is the non-representation of observational studies. Individual patient data would have provided evidence for subgroups of patients likely to benefit from EVT.

### Conclusion

In patients with LVO and low ASPECTS (3-5), EVT within 24 hrs of onset was associated with higher likelihood of achieving functional independence and early neurologic improvement but did not provide any mortality benefit in patients with large ischemic core The rates of symptomatic ICH were similar in both the groups whereas the risk of any ICH was significantly higher in the EVT arm.

## Data Availability

All data available in this manuscript can be accessed online.

## ABBREVIATIONS

ASPECTS: Alberta stroke program early CT score
CBF: Cerebral Blood Flow
CBV: Cerebral Blood Volume
DCH: Decompressive hemicraniecomy
ENI: early neurological improvement
EVT: endovascular treatment
LVO: Large Vessel Occlusion
MM: Medical Management
mRS: Modified Rankin scale score
NIHSS: National Institutes of Health Stroke Scale
NWU: Net Water Uptake
sICH: Smptomatic intracerebral haemorrhage

## References

1. Powers WJ, Rabinstein AA, Ackerson T et al; Guidelines for the Early Management of Patients With Acute Ischemic Stroke: 2019 Update to the 2018 Guidelines for the Early Management of Acute Ischemic Stroke: A Guideline for Healthcare Professionals From the American Heart Association/American Stroke Association. Stroke. 2019 Dec;50(12):e344–e418. doi: 10.1161/STR.0000000000000211. Epub 2019 Oct 30.

2. Nogueira RG, Jadhav AP, Haussen DC et al; DAWN Trial Investigators. Thrombectomy 6 to 24 Hours after Stroke with a Mismatch between Deficit and Infarct. N Engl J Med. 2018 Jan 4;378(1):11–21. doi: 10.1056/NEJMoa1706442.

3. Albers GW, Marks MP, Kemp S et al ; DEFUSE 3 Investigators. Thrombectomy for Stroke at 6 to 16 Hours with Selection by Perfusion Imaging. N Engl J Med. 2018 Feb 22;378(8):708–718. doi: 10.1056/NEJMoa1713973

4. Vahedi K, Hofmeijer J, Juettler E et al ; DECIMAL, DESTINY, and HAMLET Investigators. Early decompressive surgery in malignant infarction of the middle cerebral artery: a pooled analysis of three randomised controlled trials. Lancet Neurol. 2007;6:215–222. doi: 10.1016/S1474-4422(07)70036-4

5. Yoshimura S, Sakai N, Yamagami H et al; Endovascular Therapy for Acute Stroke with a Large Ischemic Region. N Engl J Med. 2022 Apr 7;386(14):1303–1313. doi: 10.1056/NEJMoa2118191.

6. Sarraj A, Hassan AE, Abraham MG et al ; SELECT2 Investigators. Trial of Endovascular Thrombectomy for Large Ischemic Strokes. N Engl J Med. 2023 Apr 6;388(14):1259–1271. doi: 10.1056/NEJMoa2214403.

7. Huo X, Ma G, Tong X et al ; ANGEL-ASPECT Investigators. Trial of Endovascular Therapy for Acute Ischemic Stroke with Large Infarct. N Engl J Med. 2023 Apr 6;388(14):1272–1283. doi: 10.1056/NEJMoa2213379.

8. Cagnazzo F, Derraz I, Dargazanli C et al ; Mechanical thrombectomy in patients with acute ischemic stroke and ASPECTS ≤6: a meta-analysis Journal of NeuroInterventional Surgery 2020;12:350–355

9. Diestro JDB, Dmytriw AA, Broocks G et al ; Endovascular Thrombectomy for Low ASPECTS Large Vessel Occlusion Ischemic Stroke: A Systematic Review and Meta-Analysis. Can J Neurol Sci. 2020 Sep;47(5):612–619. doi: 10.1017/cjn.2020.71. Epub 2020 Apr 17. PMID: 32299532.

10. Barber PA, Demchuk AM, Zhang J, Buchan AM. Validity and reliability of a quantitative computed tomography score in predicting outcome of hyperacute stroke before thrombolytic therapy. ASPECTS Study Group. Alberta Stroke Programme Early CT Score. Lancet. 2000;355:1670–1674. doi: 10.1016/S0140-6736(00)02237-6

11. Pexman JH, Barber PA, Hill MD, Sevick RJ, Demchuk AM, Hudon ME, Hu WY, Buchan AM. Use of the Alberta Stroke Program Early CT Score (ASPECTS) for assessing CT scans in patients with acute stroke. AJNR Am J Neuroradiol. 2001;22:1534–1542.

12. Gupta AC, Schaefer PW, Chaudhry ZA, Leslie-Mazwi TM, Chandra RV, González RG, Hirsch JA, Yoo AJ. Interobserver reliability of baseline noncontrast CT Alberta Stroke Program Early CT Score for intra-arterial stroke treatment selection. AJNR Am J Neuroradiol. 2012;33:1046–1049. doi: 10.3174/ajnr.A2942

13. Finlayson O, John V, Yeung R, Dowlatshahi D, Howard P, Zhang L, Swartz R, Aviv RI. Interobserver agreement of ASPECT score distribution for noncontrast CT, CT angiography, and CT perfusion in acute stroke. Stroke. 2013;44:234–236. doi: 10.1161/STROKEAHA.112.665208.

14. Nicholson P, Hilditch CA, Neuhaus A, Seyedsaadat SM, Benson JC, Mark I, Tsang COA, Schaafsma J, Kallmes DF, Krings T, Brinjikji W. Per-region interobserver agreement of Alberta Stroke Program Early CT Scores (ASPECTS) J Neurointerv Surg. 2020;12:1069–1071. doi: 10.1136/neurintsurg-2019-015473.

15. Berkhemer OA, Jansen IG, Beumer D, et al. Collateral status on baseline computed tomographic angiography and intra-arterial treatment effect in patients with proximal anterior circulation stroke. Stroke 2016; 47: 768–776.

16. Broocks G, Kniep H, Schramm P, et al. Patients with low Alberta Stroke Program Early CT Score (ASPECTS) but good collaterals benefit from endovascular recanalization. J Neurointerv Surg 2020; 12: 747–752.

17. Flottmann, F. et al. CT-perfusion stroke imaging: a threshold free probabilistic approach to predict infarct volume compared to traditional ischemic thresholds. Sci. Rep. 7, 6679. https://doi.org/10.1038/s41598-017-06882-w (2017).

18. McDonough, R., Elsayed, S., Meyer, L. et al. ; Low baseline ischemic water uptake is directly related to overestimation of CT perfusion-derived ischemic core volume. Sci Rep 12, 20567 (2022). https://doi.org/10.1038/s41598-022-19176-7

19. Menezes NM, Ay H, Wang Zhu M, Lopez CJ, Singhal AB, Karonen JO, Aronen HJ, Liu Y, Nuutinen J, Koroshetz WJ, Sorensen AG. The real estate factor: quantifying the impact of infarct location on stroke severity. Stroke. 2007 Jan;38(1):194–7. doi: 10.1161/01.STR.0000251792.76080.45. Epub 2006 Nov 22. PMID: 17122428.

20. Safouris A, Palaiodimou L, Szikora I, et al. Endovascular treatment for anterior circulation large-vessel occlusion ischemic stroke with low ASPECTS: a systematic review and meta-analysis. Therapeutic Advances in Neurological Disorders. 2022;15. doi:10.1177/17562864221139632

21. Broocks G, Hanning U, Flottmann F, et al. Clinical benefit of thrombectomy in stroke patients with low ASPECTS is mediated by oedema reduction. Brain 2019; 142: 1399–1407.

